# *In Vivo* Evaluation of a Biodegradable Intraanastomotic Membrane in a Porcine Model

**DOI:** 10.1101/2025.10.10.25337726

**Authors:** Daniel C. Freund, Dennis Wahl, Eberhard Grambow, Finn Jaekel, Julia Henne, Richard Kantelberg, Hans Kleemann, Friedrich Prall, Amelie R. Zitzmann, Brigitte Vollmar, Jochen Hampe, Karl Leo, Sebastian Hinz, Clemens Schafmayer

## Abstract

**Background:** Anastomotic leakage (AL) represents one of the most serious complications in gastrointestinal surgery, with reported incidence rates of up to 26 %. Despite advancements in surgical techniques, early detection of AL remains challenging, and no reliable real-time monitoring system is currently available. In this study, we investigated a resorbable polydioxanone (PDO) membrane as a potential substrate for future sensor integration, aiming to facilitate real-time monitoring of anastomotic healing.

**Methods:** In eight German Landrace pigs, 34 ileal side-to-end stapler anastomoses were examined: GM1 (n = 7), GM2 (n = 10), and controls (n = 17). Membrane stability was monitored after implantation, while adhesion formation, burst pressure, and histology were assessed on postoperative day 7.

**Results:** Both membrane geometries showed robust stability, with good anchorage of the large spokes within the anastomosis. Geometry 1 (GM1) exhibited higher burst pressure than Geometry 2 (GM2) (193 ± 43.6 vs 155 ± 65.5 mmHg, p = 0.02). Compared with controls (167 ± 42.3 mmHg), neither GM1 (p = 0.053) nor GM2 (p = 0.379) differed significantly. Adhesions occurred in all groups, without significant differences. Histological evaluation showed typical granulation tissue and fibrosis, with granulocytic inflammation more common in GM1 without affecting anastomotic stability.

**Conclusion:** This proof-of-concept study demonstrates that the PDO membrane can be safely incorporated into stapled anastomoses without compromising anastomotic healing. The membrane provides a stable, biocompatible platform suitable for future sensor integration, supporting the development of a diagnostic intraanastomotic device.

## 1 INTRODUCTION

Anastomotic leakage (AL) remains one of the most serious postoperative complications in visceral surgery, with incidence rates of up to 26 %, leading to significantly increased patient morbidity and mortality^1–3^. In addition, AL is associated with prolonged hospitalization and substantial economic burden^4^. Although early detection of anastomotic complications is critical, it remains a major clinical challenge. On average, AL is diagnosed five to eight days postoperatively^5–7^, typically based on clinical signs and nonspecific laboratory parameters, such as elevated inflammatory markers. Early diagnosis has been shown to markedly improve patient outcomes^8,9^. Currently, no reliable methods exist for early detection of AL^10,11^. The aim of this study was to develop a resorbable membrane as a potential platform for future integration of sensors, enabling real-time intraanastomotic monitoring of impaired healing, facilitating timely interventions, and potentially preventing progression to full AL. Polydioxanone (PDO) was selected for membrane fabrication due to its well-established biocompatibility and controllable hydrolytic degradation^12^. We hypothesized that incorporation of a PDO membrane into side-to-end stapled anastomoses would not compromise healing and could provide a stable, biocompatible scaffold for future sensor integration.

## 2 MATERIALS AND METHODS

### 2.1 Membrane Design

All membranes were fabricated from 150 µm thick PDO sheets (Ethicon, Inc., a Johnson & Johnson company, Somerville, NJ, USA). Following preliminary *in vitro* testing of multiple design concepts (Fig. 1A, top), two designs were selected for their favourable tissue disruption ratio, structural stability, and compatibility with the stapling device. These geometries were subsequently evaluated in the present *in vivo* study (Fig. 1A, bottom).

**Figure 1:**
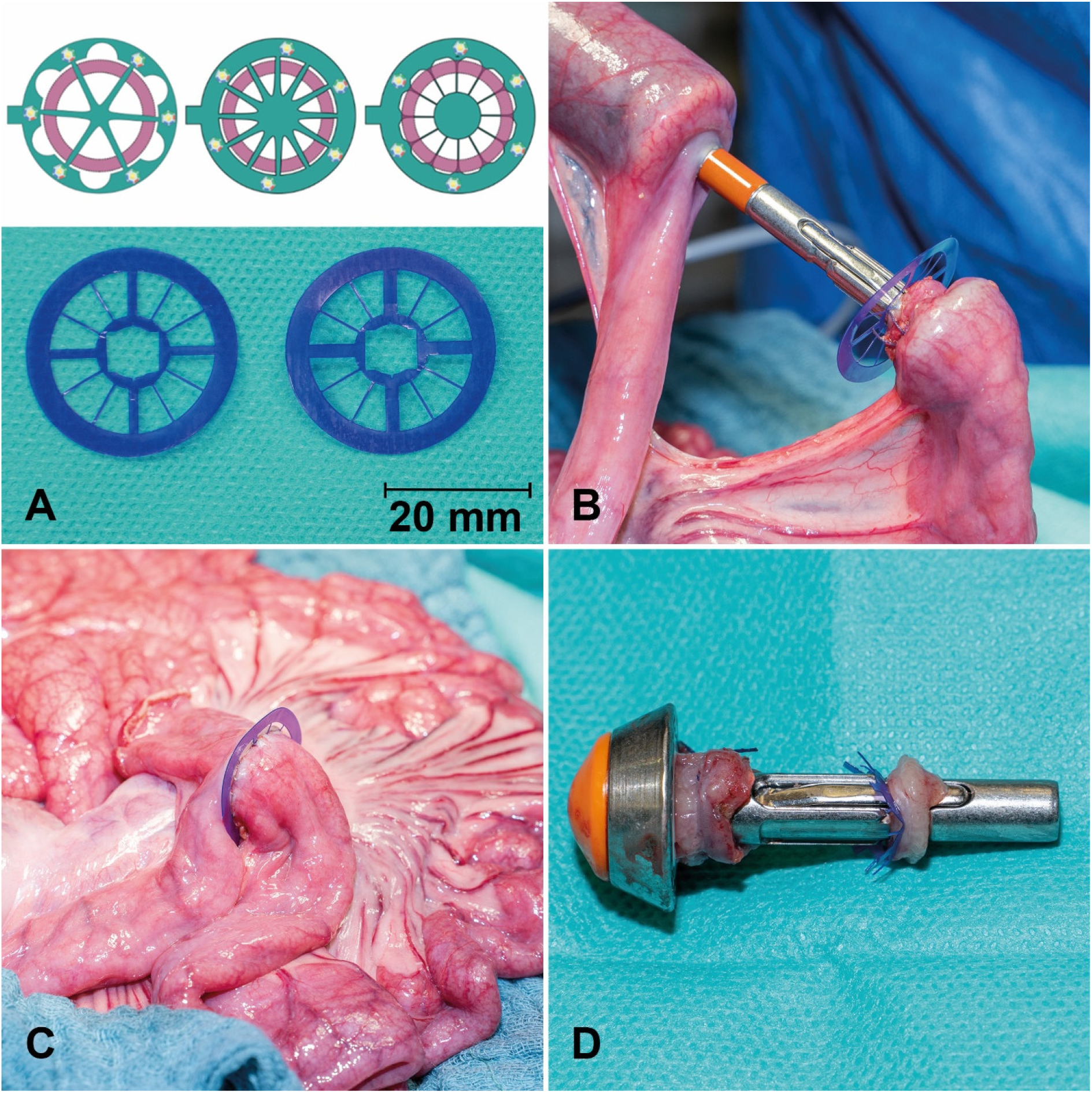
Schematic and macroscopic images of the PDO membrane with different geometries and spokes for fixation in the small bowel anastomosis. The lower photo displays GM1 (left) and GM2 (right). Each design includes four large and eight narrow spokes. **(A)**. PDO membrane before **(B)** and after implanting in a small bowel **(C)** stapler anastomosis. The anvil of the stapler with bowel margins and inner ring of the PDO membrane **(D)**.

### 2.2 Animal Model

The experimental cohort consisted of eight male German Landrace pigs, aged 12 to 16 weeks and weighing 31.6–41.0 kg (mean ± SD: 36.0 ± 3.2 kg). Another animal died prematurely due to surgical error and was excluded from the study. Post-mortem examination identified small bowel torsion proximal to the anastomotic sites. The anastomoses showed no abnormalities. Animals were housed under standardized conditions at the Central Animal Care Facility of Rostock University Medical Center and acclimatized for seven days prior to the procedure. Water was provided *ad libitum* until one day before the procedure. Premedication and anesthesia were administered according to the standard protocols of the Institute for Experimental Surgery at Rostock University Medical Center. All *in vivo* experiments were conducted in compliance with the German legislation on protection of animals (7221.3-1-050/19) and the NIH Guide for the Care and Use of Laboratory Animals (Institute of Laboratory Animal Resources, National Research Council)^14^.

### 2.3 Sedation and Anesthesia

Premedication was administered intramuscularly with 8 mg/kg azaperone (Stresnil™, Elanco, Cuxhaven, Germany), 20 mg/kg ketamine (10 % Ketamin, Medistar Arzneimittelbetrieb GmbH, Ascheberg, Germany) and 0.2 mg/kg midazolam (Dormicum, Hoffmann La Roche AG, Grenzach-Wyhlen, Germany). Animals were equipped with a pulse oximeter (Nellcor^®^ PM10N, Medtronic, Watford, UK) placed on the tail and two peripheral venous cannulas (20G, B. Braun Melsungen AG, Melsungen, Germany) in the earlobe veins. Anesthesia was induced with 200 µg fentanyl (Fentadon^®^ 50µg/ml, Eurovet Animal Health BV, Bladel, Netherlands), 100 mg propofol (Propofol 2%, MCT Fresenius, Bad Homburg, Germany) and 4 mg pancuronium (Pancuronium Inresa 4mg/2ml, Inresa Arzneimittel GmbH, Freiburg, Germany). Maintenance anesthesia was provided via total intravenous anesthesia (TIVA) with fentanyl (5-10 µg kg^-1^ h^-1^), propofol (4–8 mg kg^-1^ h^-1^), and midazolam (0.1 mg kg^-1^ h^-1^)^13^. Endotracheal intubation was performed using a 7 mm inner-diameter tube, followed by volume-controlled ventilation with a Dräger Primus^®^ ventilator, while continuously monitoring oxygen saturation, heart rate, respiratory minute volume, and end-expiratory CO_2_. Following induction, animals were placed in the supine position on the operating table and secured at all extremities.

### 2.4 Surgical Model

After sterile preparation using Braunol^®^ (B. Braun Melsungen AG, Melsungen, Germany), a midline laparotomy was performed with a careful right-sided incision around the urethra. Anastomoses were created at intervals of approximately 50 cm oral to the ileocecal valve. Side-to-end stapler anastomoses were constructed in the small intestine, allowing multiple anastomoses per animal to reduce the number of animals required for the study. After an initial mesenteric incision between the marginal arteries, the bowel was transected using monopolar cautery (ICC 300, Erbe Elektromedizin, Tübingen, Germany). The stapler anvil was inserted into the oral end of the bowel and secured with a preplaced purse-string suture (Vicryl 3.0, Ethicon^®^, Inc., a Johnson & Johnson company, Somerville, NJ, USA). The stapler (21 mm Ethicon™ Circular Stapler, Ethicon^®^, Inc., a Johnson & Johnson company, Somerville, NJ, USA) was then introduced into the aboral end, and the intestine was pierced with the trocar opposite the mesenteric side. The PDO membrane was positioned on the stapler trocar, the stapler components were connected (Fig. 1B), and the intestinal ends were approximated by gradually closing the stapler. The stapler was then fired, excising the inner membrane section to ensure anastomotic patency and prevent luminal obstruction (Fig. 1D). Following stapler removal, the blind end was resected, leaving approximately 1 cm of bowel to prevent ischemia around the anastomosis, and closed with a running suture (PDS 4-0, Ethicon^®^, Inc., a Johnson & Johnson company, Somerville, NJ, USA (Fig. 1C). In total, 34 ileal side-to-end anastomoses were examined, subdivided into three groups: GM1 (n = 7), GM2 (n = 10), and control (n = 17). Membranes were inspected for spoke displacement, and the intestines were repositioned. The abdominal wall was closed with a fascial suture (PDS sling 1.0, Ethicon^®^, Inc., a Johnson & Johnson company, Somerville, NJ, USA) and a skin staple closure (Disposable Skin Stapler F35w, ADVAN, China), followed by application of silver-aluminum spray to the wound. Depending on intraabdominal conditions, three to five anastomoses were performed per animal. Postoperatively, animals received water *ad libitum* and a standardized diet (2 × 300 g MPig-H, ssniff^®^, Soest, Germany). Oral analgesia was provided daily with 2 g metamizole (Novaminsulfon 500 mg/ml, Winthrop Arzneimittel GmbH, Frankfurt, Germany). Wounds were treated daily with iodine solution. Animal well-being was monitored using a standardized distress score (see Supplementary Material).

### 2.5 Relaparotomy, Macroscopic Evaluation and Burst Pressure Measurement

Relaparotomy was scheduled on postoperative day seven. One animal underwent relaparotomy on postoperative day five due to an elevated distress score (apathy and immobility), in accordance with the predefined study criteria. No pathological findings were detected intraoperatively, and the anastomoses remained included in the analysis. One animal underwent relaparotomy on the tenth day. The abdominal cavity was inspected for signs of complications such as peritonitis, inflammation, or bowel obstruction. The anastomoses were then identified and evaluated for macroscopic integrity and adhesion formation. Adhesions were graded according to the van der Ham score^15^ as follows: 0 = no adhesions; 1+ = minimal adhesions, primarily between the anastomosis and the omentum; 2+ = moderate adhesions, involving the omentum, anastomotic site, and adjacent small bowel loops; and 3+ = severe and extensive adhesions, including abscess formation (Fig. 2A, B). For assessment of anastomotic burst pressure, the bowel was incised 5 cm proximal and distal to the anastomosis. A catheter was inserted for isotonic saline infusion and another for pressure measurement. Both ends were securely closed, and the catheters were sealed with cable ties (Fig. 2C)^16^. The bowel segment was then continuously filled until rupture occurred, and the peak intraluminal pressure was recorded^17^. The rupture site was documented as occurring either at the anastomosis or at a distant bowel segment. Finally, while under general anesthesia, animals were euthanized with 45 mg·kg^−1^ pentobarbital (Release^®^ 300mg/ml, Wirtschaftsgenossenschaft Deutscher Tierärzte eG., Garbsen, Germany).

**Figure 2:**
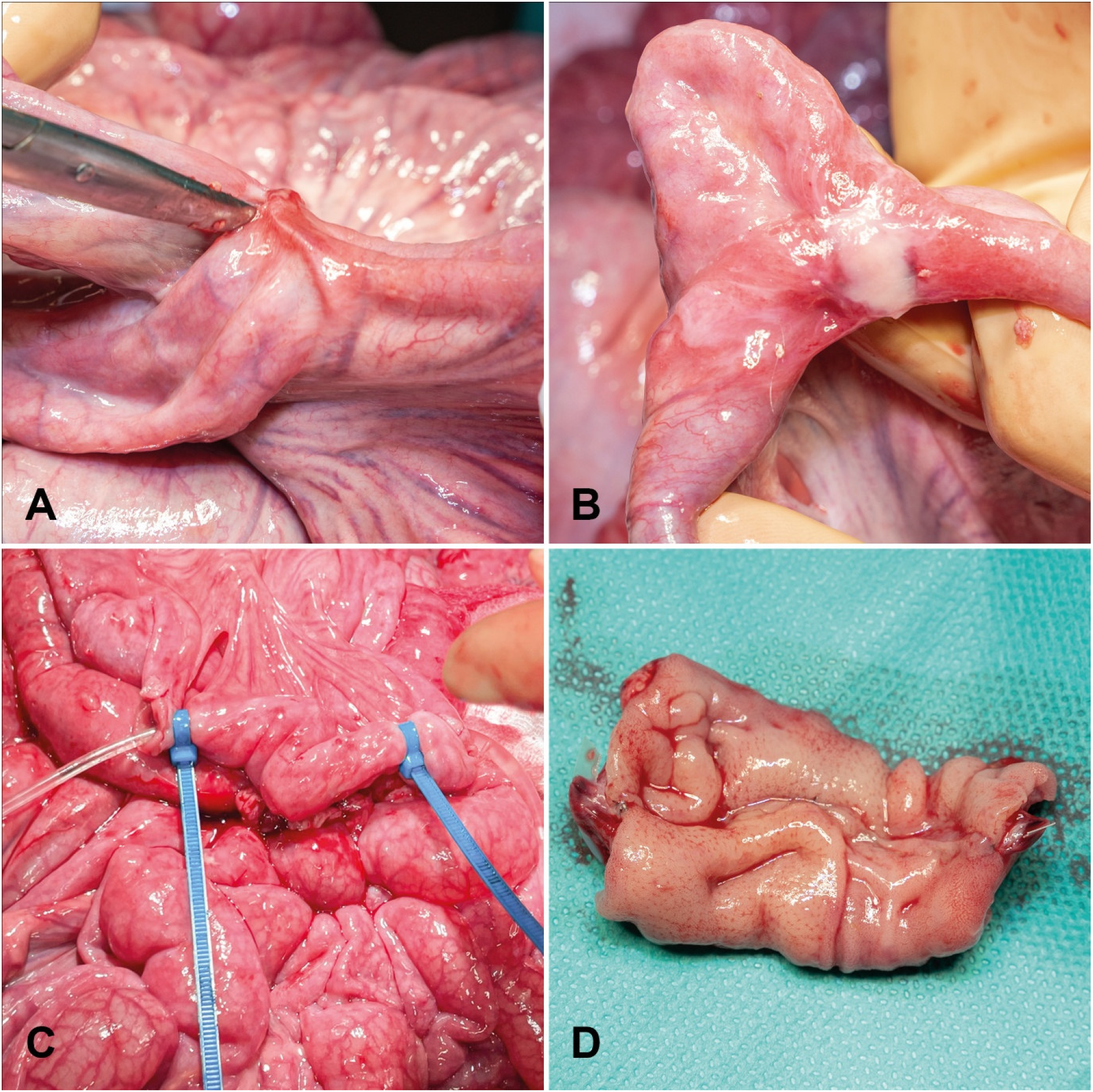
Postoperative macroscopic analysis of small bowel anastomoses on day seven. The scissors point to an adhesion (2+) **(A)**. Adhesion and fibrin deposition (1+) **(B)**. For burst pressure analysis, catheters for saline infusion (left tube) and pressure analysis (right tube) were inserted near the anastomosis and secured using cable ties **(C)**. Resected anastomosis for macroscopical study after midline incision **(D)**.

### 2.6 Histological Analysis

Intestinal segments containing the anastomoses were excised, opened along the antimesenteric border (Fig. 2D), and fixed in a stretched position in 10 % buffered formalin for 24 hours^18^. After careful removal of the metallic stapler clips, two longitudinal tissue sections, one from the mesenteric and one from the antimesenteric side, were obtained from each specimen (approximately 25 mm in length and 4 mm in thickness), with the anastomotic site positioned centrally^19^. The samples were embedded in paraffin and sectioned into 4 µm slices using a microtome (Leica RM 2145). All sections were stained with hematoxylin and eosin (H&E) and examined by a surgical pathologist blinded to the group (with or without membrane).

### 2.7 Statistical Analysis

All statistical analyses were performed using IBM SPSS Statistics for Windows, Version 29.0.2.0 (IBM Corp., Armonk, NY, USA). The Mann–Whitney U test was applied to assess differences in burst pressure between groups. The Fisher–Freeman–Halton test was used to evaluate the significance of all other data, and the Jonckheere–Terpstra test was applied to analyze the van der Ham adhesion scores. Continuous variables are presented as medians. A p-value < 0.05 was considered statistically significant.

## 3 RESULTS

### 3.1 Membrane Design and Stability

After initial design optimization, two membrane prototypes were evaluated *in vivo*. Both designs comprised an outer ring (28.88 mm outer diameter, 21.38 mm inner diameter) connected to an inner ring (10 mm outer diameter) by eight narrow and four wide spokes (Fig. 1A). GM1 featured 1.5 mm-wide spokes, while GM2 incorporated 2.0 mm-wide spokes; both designs included 0.6 mm narrow spokes. A central circular cutout (6.4 mm) accommodated the trocar of a 21 mm circular stapler, with additional peripheral cuts aligned to the trocar’s widest points to prevent rotation or dislocation during implantation. The spoke configuration was designed to maximize surface area for future sensor integration while maintaining stable anchoring and minimizing the amount of material embedded within the intestinal wall to reduce interference with anastomotic healing. After stapled implantation, membranes were examined for dislocation or structural damage (Fig. 3A). Outer ring damage was observed in one GM1 anastomosis. Dislocation of large spokes was infrequent (GM1: 14.3 %; GM2: 20.0 %; p = 0.640; Fig. 3A). Small spoke dislocation occurred in both geometries: in GM1, 43.9 % of anastomoses showed all small spokes intact, none exhibited single spoke dislocation, and 57.1 % showed dislocation of two spokes; in GM2, 40.0 % had all spokes intact, 30.0 % exhibited dislocation of one spoke, and 30.0 % showed dislocation of two spokes. There was no statistically significant difference in the rate or extent of small spoke dislocation between groups (p = 0.358).

**Figure 3:**
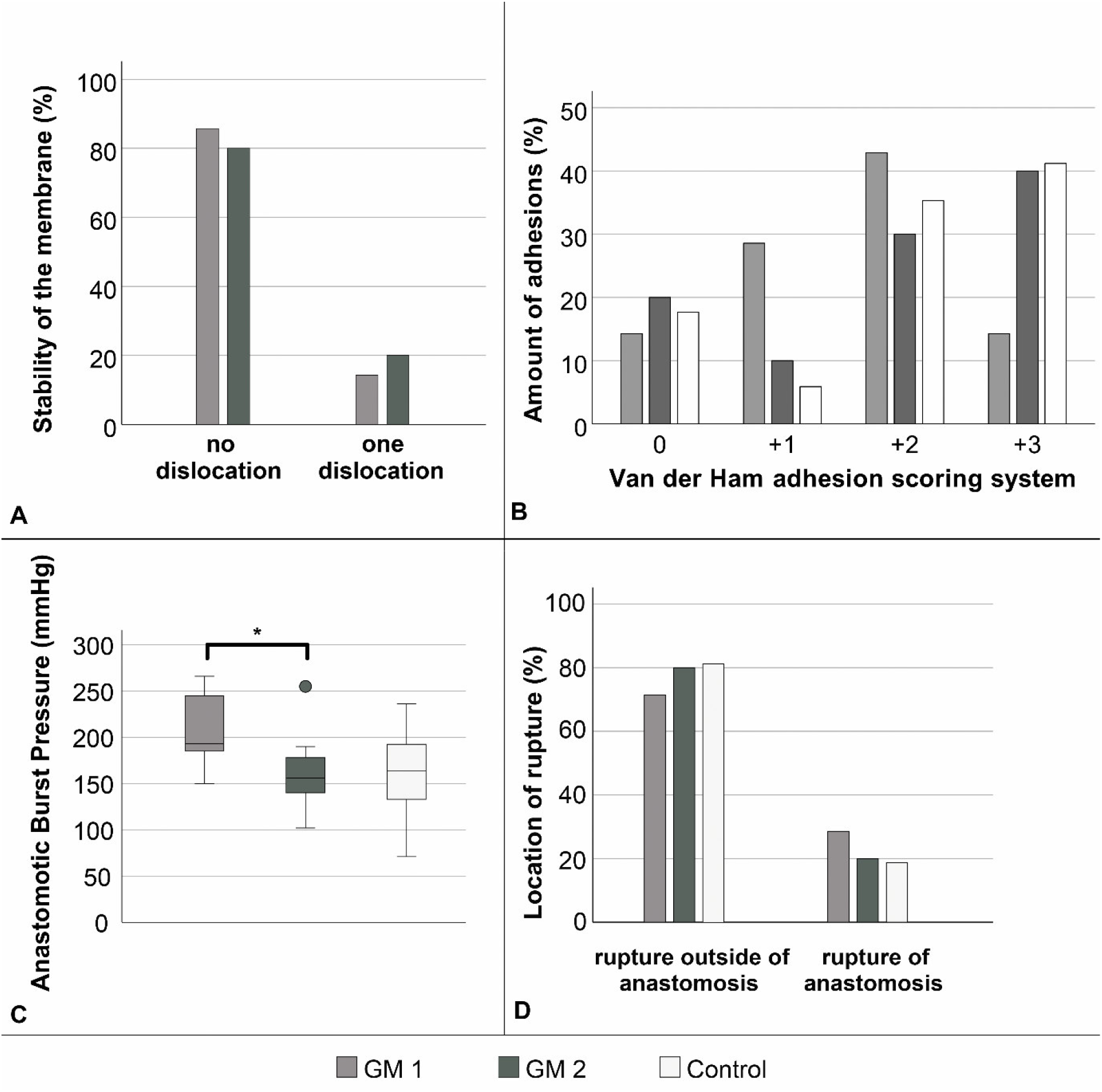
Quantitative analysis of PDO membrane stability immediately after implantation in a small bowel stapler anastomosis. Stability was assessed by dislocation large spokes of the membrane. No significant differences between the groups (*p* = 0.640; Fisher–Freeman–Halton test) **(A)**. Analysis of anastomotic stability. Quantitative assessment of Van der Ham adhesion score based on adhesion formation around the anastomosis showed no significant difference (*p* = 0.358; Jonckheere–Terpstra test) **(B)**. Stability was further studied by means of burst pressure; the difference between GM1 and GM2 was significant (*p* = 0.02; Mann–Whitney U test), while no significant differences were observed compared with the control group (GM1 vs Control: *p* = 0.053; GM2 vs Control: *p* = 0.379) **(C)**. The location of rupture during burst pressure assessment showed no significant difference (*p* > 0.05; Fisher–Freeman–Halton test) **(D)**. GM1: n = 7, GM2: n = 10, Control: n = 17.

### 3.2 Macroscopic Examination

No AL or stenosis was observed in any group. All anastomoses were patent and free of stenosis. Adhesions were present in all groups, with variable severity (Fig. 2A,B; 3B). In GM1, 14.3 % of anastomoses showed no adhesions, 28.6 % had mild adhesions (+1), 42.9 % moderate (+2), and 14.3 % severe (+3). In GM2, 20 % were adhesion-free, 10 % mild, 30 % moderate, and 40 % severe. In the control group, 17.6 % showed no adhesions, 5.9 % mild, 35.3 % moderate, and 41.2 % severe. Mild adhesions were more frequent in GM1, whereas severe adhesions predominated in GM2 and controls. Overall, differences in adhesion distribution among groups were not statistically significant (p = 0.358).

### 3.3 Anastomotic Burst Pressure

Anastomotic burst pressure was highest in GM1 (193 ± 43.6 mmHg) and lowest in GM2 (155 ± 65.5 mmHg), compared with 167 ± 42.3 mmHg in the control group (Fig. 3C). The difference between GM1 and GM2 reached statistical significance (p = 0.02), whereas comparisons of GM1 or GM2 with controls were not significant (p = 0.053 and p = 0.379, respectively). In most cases, intestinal rupture occurred outside the anastomosis (71.4 % in GM1, 80 % in GM2, and 82.4 % in controls; p = 0.861; Fig. 3D), with no notable differences between groups.

### 3.4 Histological Examination

78 specimens were submitted to histological examination and the anastomoses were visualized in 77 of these. Granulation tissue and fibroblast-rich fibrosis were found in all anastomoses, frequently in zonal arrangements and often surrounding residual surgical material (Fig. 4 A, C). However, differences between anastomoses were observed as follows and scored in a systematic slide review as present or absent: granulocytic inflammation adjacent to surgical material **(GIS)** (Fig. 4 B, D); abscess formation independent of residual surgical material **(IAF)**; surgical induced mucosal hernias without abscess formation **(HAS)**, abscess formation in surgically induced mucosal hernias **(HWA)**; purulent exudate on the peritoneum **(PEP)**. The slide review was done blinded to anastomosis types and the type of membrane (GM1 versus GM2). The results of the two slices per anastomosis were combined into one result, basing the final score on the more severe histological finding. GIS was found more frequently in the groups of the membranes, especially in group GM1 (GM1: 42.9 %, GM2: 30.0 % Control: 11.8 %; p = 0.221). PEP also occurred most frequently in GM1 and was least frequent in the control group (GM1: 42.9 %, GM2: 10.0 %, Control: 5.9 %; p = 0.076). In contrast, HWA was found exclusively in the control group (Table 1). However, the differences between the three groups were not statistically significant.

**Table 1:** Results of the histological examination: granulocytic inflammation adjacent to surgical material **(GIS)**; abscess formation independent of residual surgical material **(IAF)**; surgical induced mucosal hernias without abscess formation **(HAS)**, abscess formation in surgically induced mucosal hernias **(HWA)**; purulent exudate on the peritoneum **(PEP)**. The difference between the groups does not reach statistical significance (all p-values > 0.05; Fisher-Freeman-Halton test).

|  | GM 1 Group<br>in (%) | GM 2 Group<br>in (%) | Control Group<br>in (%) | p-value |
| --- | --- | --- | --- | --- |
| **GIS** | 42.9 | 30.0 | 11.8 | 0.221 |
| **IAF** | 14.3 | 10.0 | 5.9 | 0.773 |
| **HAS** | 0 | 20.0 | 5.9 | 0.415 |
| **HWA** | 0 | 0 | 11.8 | 0.697 |
| **PEP** | 42.9 | 10.0 | 5.9 | 0.076 |

**Figure 4:**
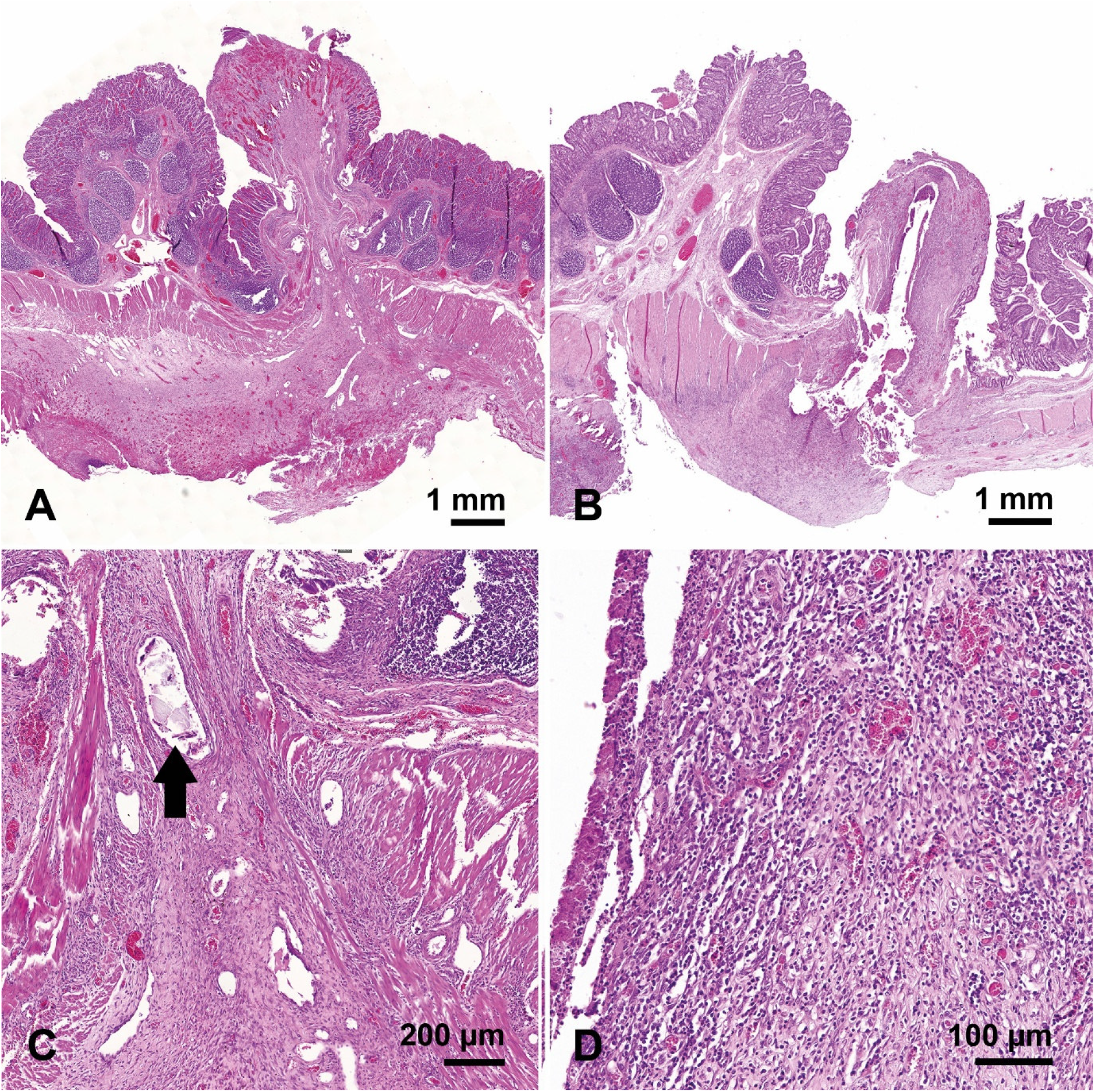
Microscopic images of two anastomoses, both from group GM2: **(A)** Panoramic view of an anastomosis without significant granulocytic inflammation; **(C)** at higher magnification residual PDO substrate is seen (arrow) which is surrounded by a fibrosing reaction. **(B)** Panoramic view of an anastomosis with substantial granulocytic inflammation (scored GIS) around residual PDO substrate, which at higher magnification **(D)** is fully appreciated.

## 4 DISCUSSION

### 4.1 Principal Findings

This study suggests that integration of a PDO membrane into stapled anastomoses does not impair anastomotic healing, supporting its potential as an intraanastomotic carrier membrane for sensor devices. GM1 exhibited slightly higher macroscopic healing performance than GM2, although this difference reached statistical significance only in burst pressure measurements. No group performed worse than comparable data reported in the literature^16^. All other macroscopic parameters were similar across the groups. Moderate to severe adhesions were observed in all groups. This design was deliberately chosen to minimize total animal use in accordance with the 3R principle. Comparable adhesion scores and the absence of significant intergroup differences suggest that the membrane itself did not substantially contribute to adhesion formation. Histological analysis indicated a higher frequency of granulocytic inflammation and purulent exudate in GM1 compared with GM2 (Table 1), suggesting a modest local tissue response. However, these differences were not statistically significant and likely reflect a physiological reaction to resorbable material, without impacting anastomotic stability. PDO has a long history of clinical use as a suture and implant material, with well-established biocompatibility and predictable *in vivo* degradation kinetics^12^. This study confirms the mechanical stability of the membrane and its compatibility with standard surgical techniques. The membrane could be reliably introduced through the circular stapler without technical difficulty, and the stapler functioned normally with the membrane in place. Large spokes remained stable, supporting potential future circumferential sensing. The outer ring was largely intact, with a single break in GM2 resulting from moisture-related accelerated PDO^12^ degradation due to non-airtight storage, which could be prevented with improved industrial packaging. Dislocation of small spokes occurred frequently but was not considered critical, as they primarily support implantation and are not intended for sensor integration.

### 4.2 Clinical Relevance and Translational Outlook

AL is influenced by multiple factors, some modifiable intraoperatively (e.g., blood loss, fecal contamination, operative time), and others patient-specific (e.g., chronic kidney disease, diabetes, hypertension, smoking), which are prognostic for AL^20–24^. These factors often affect microvascular perfusion and wound healing, meaning not all AL can be prevented. Non– technically induced insufficiencies typically develop gradually, offering an opportunity for early detection prior to perforation or peritonitis. Currently, no clinical device provides real-time, direct monitoring of anastomotic healing. Radiologic methods, such as CT imaging, have limited sensitivity, as shown by Doeksen et al.^25,26^, and involve radiation exposure that should be avoided without clinical indication^26,27^. Endoscopic examination provides sufficient sensitivity and specificity but is invasive. Recent studies have explored bioresorbable sensors, including impedance-based systems^28^, magnesium electrodes^29^, and wireless oxygen or pH sensors^30^. However, these approaches generally provide only single-point measurements or require external positioning, limiting circumferential coverage. In contrast, the PDO membrane provides a mechanically stable, circumferential platform for potential site-specific sensing along the entire staple line. At the same time, several studies have already shown that impedance measurements can be used to recognize anastomotic insufficiencies at an early stage^31,32^. Previous *ex vivo* work demonstrated the feasibility of integrating resorbable electronic elements directly onto PDO membranes using screen-printable zinc and silver inks, confirming mechanical stability, biocompatibility, and controlled degradation^33^. Future studies will focus on incorporating sensors (impedance-, oxygen-, or lactate-based^34–36^) onto the membrane surface to enable continuous, localized monitoring of anastomotic healing. The circumferential design may allow early detection of impaired healing, supporting timely intervention. From a translational perspective, the use of clinically established PDO and standard manufacturing techniques such as extrusion or thermoforming enables scalable, cost-efficient production of sensor-integrated membranes compatible with existing medical-grade polymers once sensor functionality and long-term safety have been validated.

This study demonstrates that a bioresorbable PDO membrane can be safely integrated into stapler anastomoses in a porcine model without compromising anastomotic healing. The membrane provides a mechanically stable, biocompatible platform suitable for future development of smart anastomotic devices.

### 4.3 Limitations

The obtained results are part of a proof-of-concept study; therefore, the sample size was not calculated by a power-analysis. To finally evaluate the influence on long-term anastomotic healing, a power analysis-based case-control study should be performed including extended clinical and histological follow-up. Consequently, the translational potential to human application remains preliminary. Future studies should address sensor material integration and prolonged observation periods.

## 5 Conclusion

This proof-of-concept study demonstrates the technical feasibility of integrating a resorbable PDO membrane into stapled small bowel anastomoses without compromising anastomotic integrity, thereby highlighting its potential for future sensor integration. The results are encouraging, but further studies with larger cohorts and extended follow-up are required to confirm safety beyond a reasonable doubt.

## Supporting information

Score_Sheet

## Data Availability

All data produced in the present study are available upon reasonable request to the authors

## Acknowledgment

We thank the staff of the Rudolf Zenker Institute for Experimental Surgery for their excellent animal care and valuable assistance in preparing the histological sections. We also gratefully acknowledge Ms. Burmeister for her support with graphic design and for taking photographs. RK thanks the Hector Fellow Academy for Funding and Support.

## Conflict of Interest

Authors D.W., F.J., D.F., E.G., H.K., J.H., S.H., C.S., K.L. declare to be inventors on patent number DE 10 2024 137 023.8, which is related to this script. Other authors declare no conflict of interest.

## Fundings

This project is funded by the DFG – German Research Foundation under project number 461264398 and grant number SCHA 1686/6-1. The PDO-membrane substrate was provided free of charge by Ethicon^®^, Inc., a Johnson & Johnson company, Somerville, NJ, USA.

