## Supplementary material for "*In Vivo* Evaluation of a Biodegradable Intraanastomotic Membrane in a Porcine Model": Score_Sheet

### Score Sheet and Endpoints of the Study

**Score Sheet (Observation Interval: 1 time per day)**

| **I Observation** | **Score** |
|---|---|
| Hemodynamic instability with persistent need for vasopressors | 1 |
| Hypoxia with $SpO_2$ saturation below 50% lasting more than 5 minutes | 1 |
| Persistent uncontrollable bleeding | 3 |
| **II. General Conditions** | **Score*** |
| Scaly skin; cloudy eyes | 1* |
| Clogged body openings; abnormal posture; dehydration | 1* |
| Seizures, paralysis, abnormal respiratory sounds, hypothermia | 2* |
| **III. Spontaneous Behavior** | **Score*** |
| Isolation | 1* |
| Apathy (almost no voluntary movement observable) | 2* |
| Signs of pain | 3* |
| Automutilation | 3* |
| **IV. Procedure-Specific Criteria** | **Score*** |
| Open wounds | 1* |
| **Total Theoretical Score** | **0-19** |

**Actions / Endpoints of Study**

| Single Score | Total Score | Burden | Action |
|---|---|---|---|
| * | - | - | The trial did not start |
| 1 (IV) | 1 | low | Wound revision under anesthesia |
| 3 (III) | 3-9 | severe | Termination of trial via administration of pentobarbital or re-laparotomy under anesthesia |
| 1-2 (III) | 1-3 | moderate | Clinical re-evaluation of spontaneous behavior after 60 min; if unchanged, anesthesia and re-laparotomy |
| 2 | 2-4 | severe | Animals are anesthetized and undergo re-laparotomy prior to euthanasia |
| 1 | 1-2 | low | Clinical re-evaluation after 4h; if unchanged, anesthesia and re-lap and possibly euthanasia |
| 1 (I) | 1 | low | Optimization of volume management, possibly revision of surgical site |
| 3 (I) | 3-5 | severe | The trial is terminated by administration of pentobarbital |
